# How Large Language Models Perform on the United States Medical Licensing Examination: A Systematic Review

**DOI:** 10.1101/2023.09.03.23294842

**Authors:** Dana Brin, Vera Sorin, Eli Konen, Girish Nadkarni, Benjamin S Glicksberg, Eyal Klang

**Affiliations:** Department of Diagnostic Imaging, Chaim Sheba Medical Center, Tel Hashomer, Israel; The Faculty of Medicine, Tel-Aviv University, Israel; DeepVision Lab, Chaim Sheba Medical Center, Tel Hashomer, Israel; Division of Data-Driven and Digital Medicine (D3M), Icahn School of Medicine at Mount Sinai, New York, New York, USA; The Charles Bronfman Institute of Personalized Medicine, Icahn School of Medicine at Mount Sinai, New York, New York, USA; Hasso Plattner Institute for Digital Health, Icahn School of Medicine at Mount Sinai, New York, New York, USA

## Abstract

**Objective:** The United States Medical Licensing Examination (USMLE) assesses physicians’ competency and passing is a requirement to practice medicine in the U.S. With the emergence of large language models (LLMs) like ChatGPT and GPT-4, understanding their performance on these exams illuminates their potential in medical education and healthcare.

**Materials and Methods:** A literature search following the 2020 PRISMA guidelines was conducted, focusing on studies using official USMLE questions and publicly available LLMs.

**Results:** Three relevant studies were found, with GPT-4 showcasing the highest accuracy rates of 80-90% on the USMLE. Open-ended prompts typically outperformed multiple-choice ones, with 5-shot prompting slightly edging out zero-shot.

**Conclusion:** LLMs, especially GPT-4, display proficiency in tackling USMLE-standard questions. While the USMLE is a structured evaluation tool, it may not fully capture the expansive capabilities and limitations of LLMs in medical scenarios. As AI integrates further into healthcare, ongoing assessments against trusted benchmarks are essential.

## INTRODUCTION

The United States Medical Licensing Examination (USMLE) is a multi-part professional exam that is mandatory to practice medicine in the United States. It is a rigorous assessment of physicians’ knowledge and skills, providing a standardized measure of competence for both domestic and international medical graduates.(1–4) As such, the USMLE has become a critical benchmark for medical education and is increasingly used in research as a standard for testing the capabilities of various healthcare-focused artificial intelligence tools.

Advancements in natural language processing (NLP), have led to the development of large language models (LLMs) like GPT-3 and GPT-4. These models can generate human-like text and perform various complex NLP tasks. LLMs are increasingly studied in healthcare for different applications, including aiding diagnosis, streamlining administrative tasks, and enhancing medical education.(5–8) It is critical to assess the performance of these models in a standardized and rigorous manner, especially in specialized fields like medicine.(6,9)

Given the important role of the USMLE in assessing medical competence, understanding LLMs fare on this test offers valuable insights into their clinical reasoning, potential applications, and limitations in healthcare. Thus, the aim of our study was to systematically review the literature on the performance of publicly available LLMs on official USMLE questions, analyze their clinical reasoning capabilities, and determine the impact of various prompting methodologies on outcomes.

## METHODS

### Literature search

A systematic literature search was conducted for studies on LLM’s performance on the USMLE.

We searched for articles published up to July 2023. PubMed and Google Scholar were used as databases. Search keywords included “USMLE”, “United Stated Medical License Exams”, “LLM” and “Large Language Models”. We also searched the references lists of relevant studies for any additional relevant studies. **Figure 1** presents a flow diagram of the screening and inclusion process.

**Figure 1.**
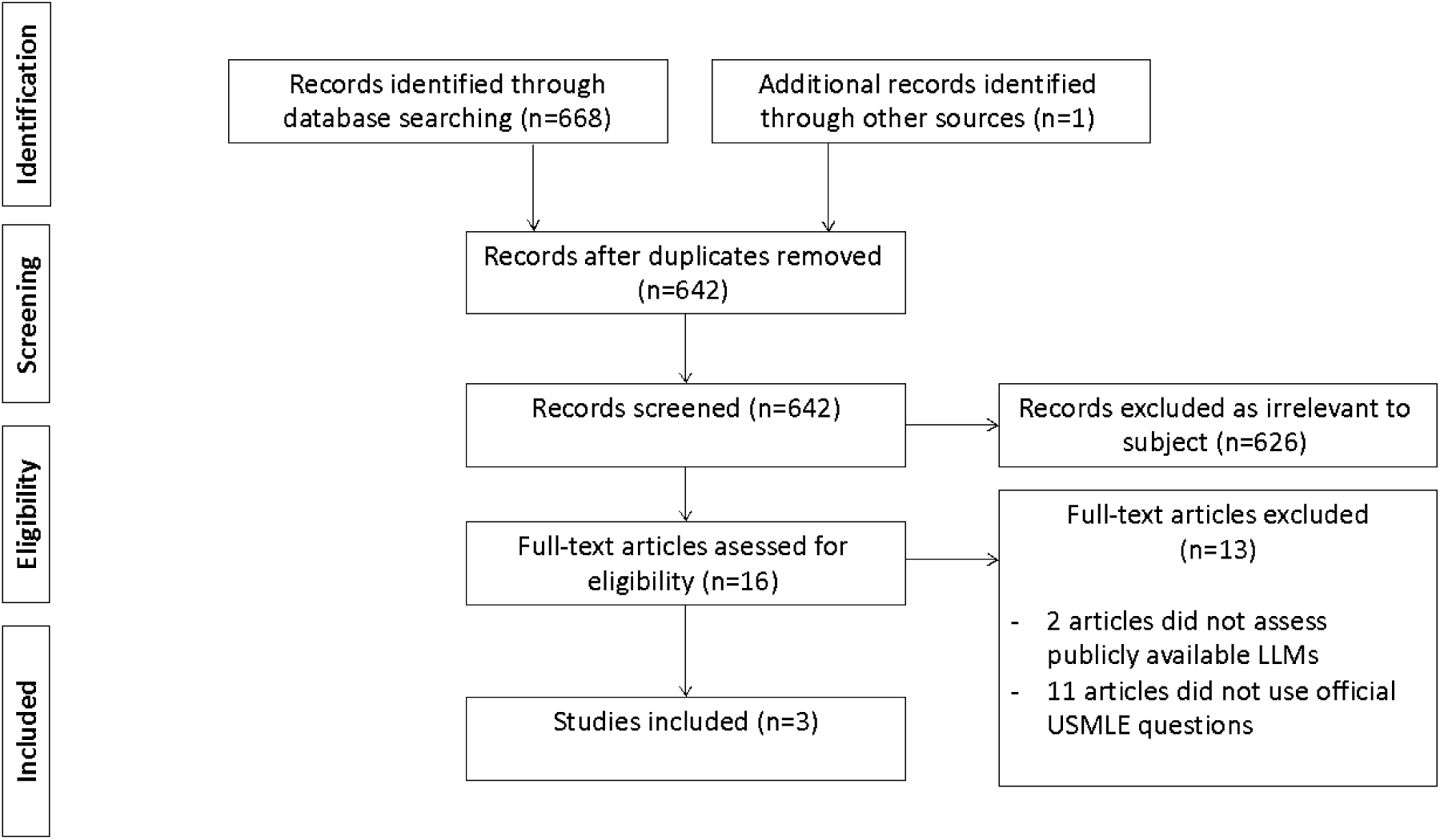
Flow diagram of the search and inclusion process in the study. The study followed the Preferred Reporting Items for Systematic Review and Meta-Analyses (PRISMA) guidelines. USMLE = United States Medical Licensing Examination. LLMs = Large Language Models.

### Eligibility Criteria

We included studies that evaluated the performance of publicly available LLM’s on official USMLE questions. We excluded papers that evaluate LLMs using unofficial sources of USMLE-like questions (e.g., MedQA) and non-English papers.

### Screening and Synthesis

This review was reported according to the 2020 Preferred Reporting Items for Systematic Reviews and Meta-Analysis (PRISMA) guidelines.(10)

## RESULTS

Data were extracted from three publications that evaluated the performance of publicly available LLMs on USMLE questions. The parameters evaluated in each publication are described in **Table 1**.

**Table 1.** Publications reporting on performance of LLMs in USMLE questions.

| Study<br>(Ref) | Month,<br>Year | Journal | Model(s)<br>tested | USMLE<br>step(s)<br>evaluate<br>d | Questions<br>sets used | Prompting<br>Methodology | Images<br>and<br>graphs | Performance<br>metric(s) used |
| --- | --- | --- | --- | --- | --- | --- | --- | --- |
| Kung et<br>al.(18) | February<br>2023 | PLOS<br>digital<br>health | ChatGPT | Step<br>1,2,3 | Sample<br>exam* | Open-ended,<br>Multiple<br>choice,<br>Multiple<br>choice with<br>forced justification | Excluded | Accuracy, concordance,<br>and insight |
| Glison<br>et<br>al.(14) | August<br>2023 | JMIR<br>medical<br>education | ChatGPT,<br>GPT-3,<br>InstructGP<br>T | Step 1,2 | Sample<br>exam* | Multiple<br>choice | Excluded | Logical justification,<br>presence of information<br>internal to the question,<br>presence of information<br>external to the question |
| Nori et<br>al.(17) | April 2023 | arXiv | GPT-4,<br>GPT-3.5 | Step<br>1,2,3 | Sample<br>exam*,<br>Self-<br>assessme<br>nt** | Open ended<br>template:<br>zero-shot,<br>5-shot | Included | - |

\* Sourced from the official USMLE website
\*\* Sourced from the official NMBE website

### Data sets

There are two official sources for USMLE questions – USMLE Sample exam, which is freely available,(11) and the NMBE Self-Assessment, available for purchase at the NMBE website.(12) Both include questions for Step 1, 2CK and 3.

### Large Language Models

The large language models included in this study were all developed by OpenAI.(13) GPT-3 is an autoregressive model known for its ability to handle a variety of language tasks without extensive fine-tuning. GPT-3.5, a subsequent version, serves as the foundation for both ChatGPT and InstructGPT. ChatGPT is tailored to generate dialogic responses across diverse topics, while InstructGPT is designed to provide detailed answers to specific user prompts. Both models, although sharing a foundational architecture, have been fine-tuned using different methodologies and datasets to cater to their respective purposes.

GPT-4, though specifics are not fully disclosed, is recognized to have a larger scale than its predecessor GPT-3.5, indicating improvements in model parameters and training data scope.(14–16)

When evaluated on USMLE questions, GPT-4 outperformed all other models with accuracy rates of 80-86%, 81-89% and 81-90% in Step1, Step2 and Step3, respectively. ChatGPT also had relatively good results, outperforming GPT-3, InstructGPT and GPT-3.5 with accuracy rates of 41-75%, 49-61% and 55-68% in Step1, Step2 and Step3, respectively (**Tables 2-4**).

**Table 2.** LLM’s performance (%) on USMLE Step1.

| Ref. | Data Set | Prompting | GPT-3 | InstructGP<br>T | GPT-3.5 | ChatGPT | GPT-4 |
| --- | --- | --- | --- | --- | --- | --- | --- |
| (18) | Sample exam | Open-ended<br>Multiple choice<br>Multiple choice with forced justification |  |  |  | 75/45.4*<br>55.8/36.1*<br>64.5/41.2* |  |
| (14) | Sample exam | Multiple choice | 25.3 | 51.7 |  | 64.4 |  |
| (17) | Self-assessment | Zero-shot<br>5-shot |  |  | 49.62<br>54.22 |  | 83.46<br>85.21 |
|  | Sample exam | Zero-shot<br>5-shot |  |  | 51.26<br>52.10 |  | 80.67<br>85.71 |

\* With indeterminate responses included

**Table 3.**
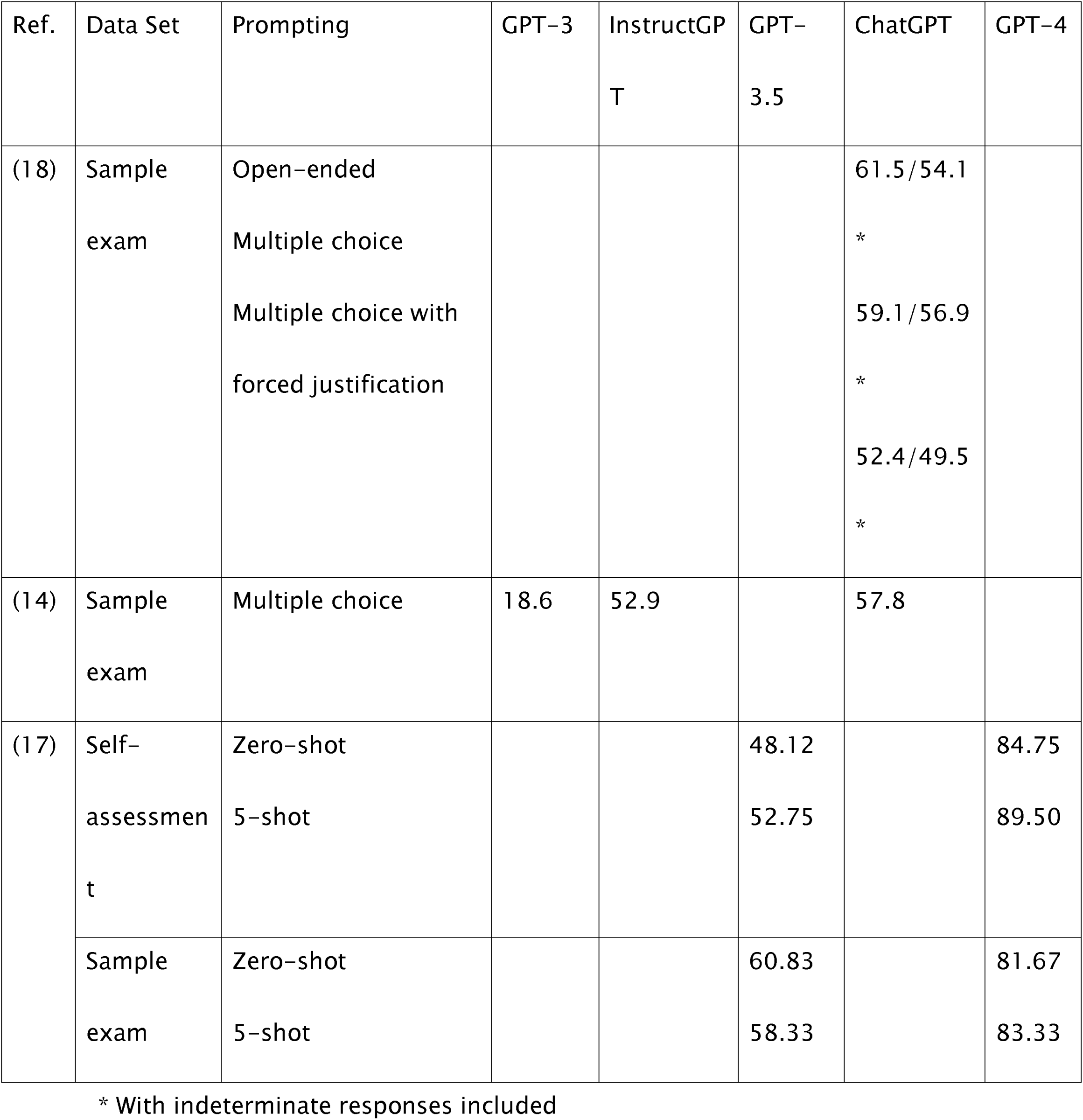
LLM’s performance (%) on USMLE Step2.

**Table 4.**
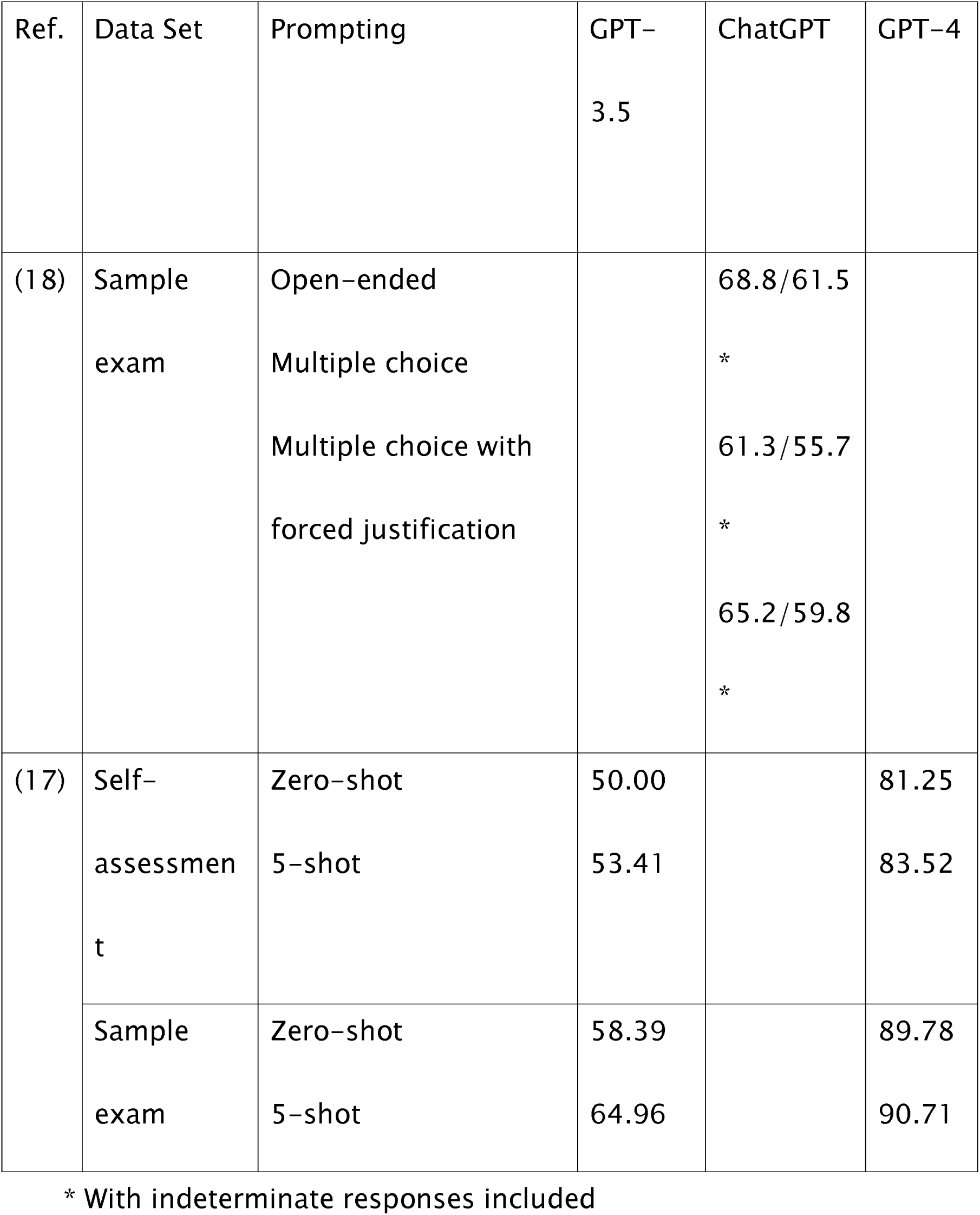
LLM’s performance (%) on USMLE Step3.

### Questions with Media Elements

Some of the USMLE questions use media elements such as graphs, images, and charts (14.4% and 13% of questions in the Self-assessment and Sample exam, accordingly).(17) While Glison et al.(14) and Kung et al.(18) excluded these questions because these elements do not get passed to the model, Nori et al.(17) included them in the evaluation and found that while GPT-4 performs best on text-only questions, it still performs well on questions with media elements, with 68-79% accuracy, despite not being able to see the relevant images. GPT-3.5 was found to have a 41-53% accuracy in the media elements containing questions.

### Prompting methods

Different prompting methods were used to test the performance of the LLMs. Examples of prompts are shown in **Table 5**.

**Table 5.**
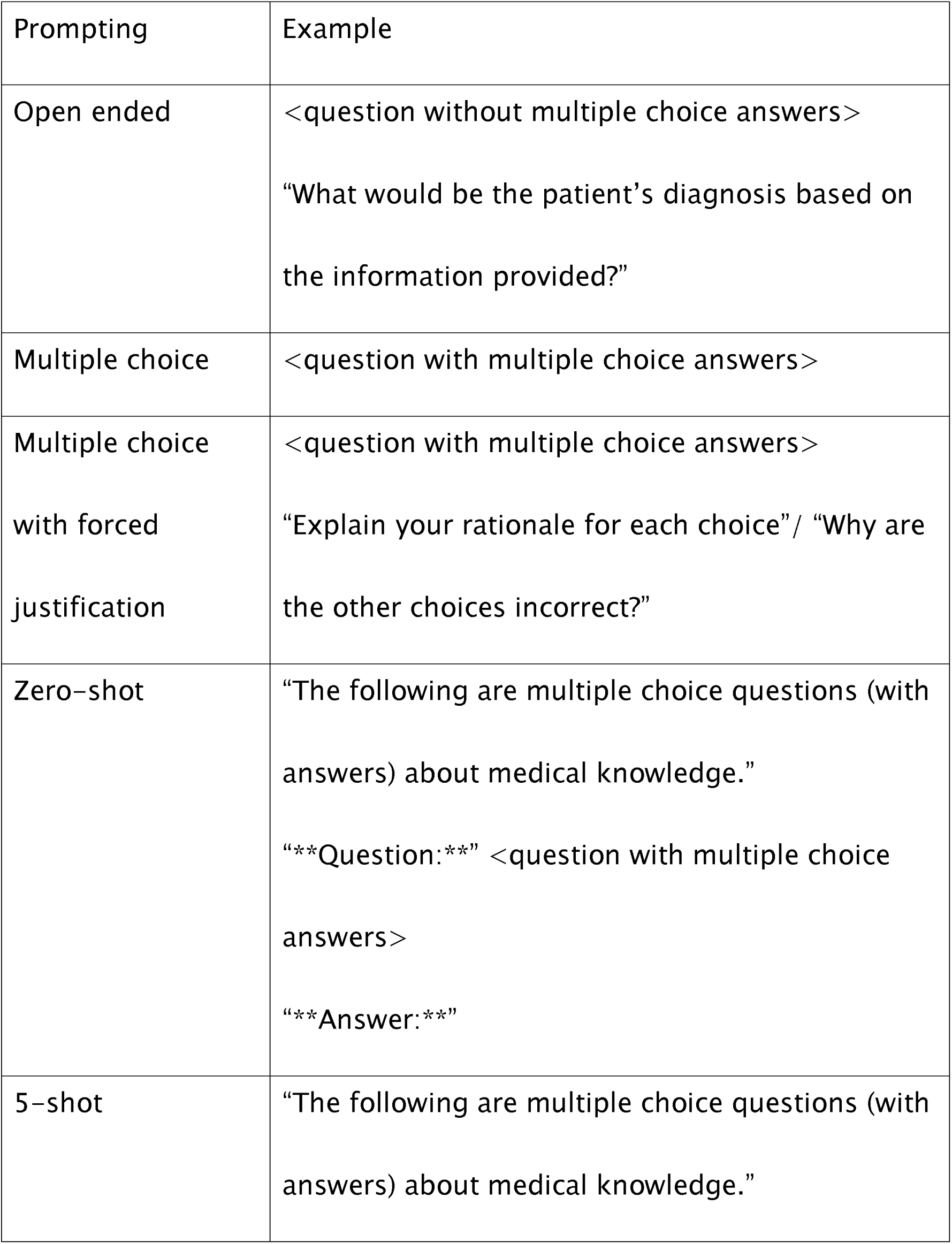
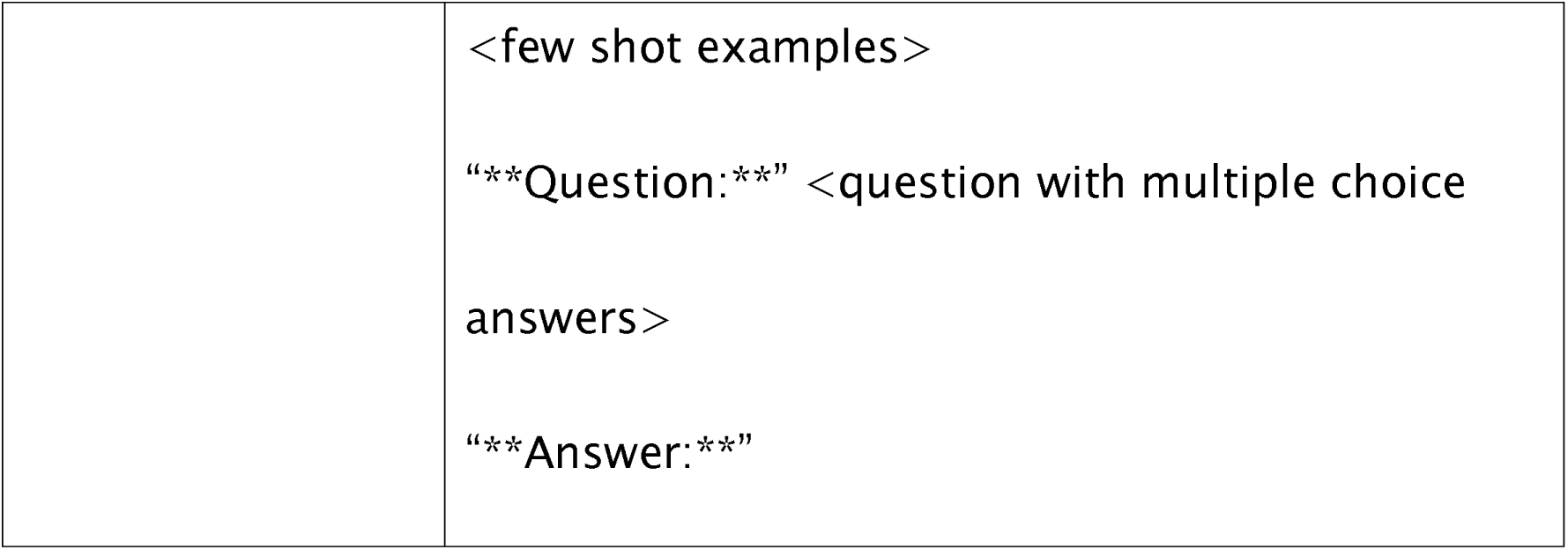
Prompt templates used to assess USMLE questions. Elements between <> are replaced with question-specific data.

| Prompting | Example |
| --- | --- |
| Open ended | <question without multiple choice answers><br>“What would be the patient’s diagnosis based on the information provided?” |
| Multiple choice | <question with multiple choice answers> |
| Multiple choice with forced justification | <question with multiple choice answers><br>“Explain your rationale for each choice”/ “Why are the other choices incorrect?” |
| Zero-shot | “The following are multiple choice questions (with answers) about medical knowledge.”<br>“**Question:**” <question with multiple choice answers><br>“**Answer:**” |
| 5-shot | “The following are multiple choice questions (with answers) about medical knowledge.” |
|  | <few shot examples><br>“**Question:**” <question with multiple choice<br>answers><br>“**Answer:**” |

Kung et al. (18) tested three prompting options. In the Open-Ended format, answer choices were eliminated, and variable lead-in interrogative phrases were incorporated to mirror natural user queries. In the Multiple-Choice Single Answer without Forced Justification format, USMLE questions were reproduced exactly. The third format, Multiple Choice Single Answer with Forced Justification, required ChatGPT to provide rationales for each answer choice.

Glison et al.(14) used only multiple choice prompting.

Nori et al. (17) also used multiple choice questions but tested both zero-shot and few-shot prompting. Zero-shot prompting requires a model to complete tasks without any prior examples, utilizing only the knowledge gained from pre-training. On the other hand, few-shot prompting provides the model with limited examples of the task at hand before execution. The model is expected to generalize from these examples and accurately perform the task.

Overall, the performance of the models was slightly affected by the prompting method, with open-ended prompting showing better results than multiple choice, and 5-shot prompting giving better results than zero-shot.

### Performance assessment

All papers assessed accuracy of the models in answering USMLE questions. Two studies also included qualitative assessment of the answers and explanations provided by the LLMs. Glison et al.(14) assessed each answer for logical reasoning (identification of the logic in the answer selection), use of information internal to the question, and use of information external to the question. ChatGPT was reported to use information internal to the question in 97% of questions. The use of information external to the question was higher in correct (90-93%) answers than incorrect answers (48-63%). In addition, every incorrect answer was labeled for the reason of the error: logical error (the response uses the pertinent information but does not translate it to the correct answer), information error (did not identify the key information needed) or statistical error (an arithmetic mistake). Logical errors were the most common, found in 42% of incorrect answers. (14) Kung et al.(18) evaluated each output for concordance and insight, by two physician reviewers. A high concordance of 94.6% was found across ChatGPT’s answers. ChatGPT produced at least one significant insight in 88.9% of questions. The density of insight contained within the explanations provided by ChatGPT was significantly higher in questions answered accurately than in incorrect answers.

## DISCUSSION

This review provides a comparative analysis of LLMs performance on USMLE questions. While GPT-4 secured accuracy rates within the 80-90% range, ChatGPT demonstrated competent results, outpacing the capabilities of previous models, GPT-3, InstructGPT, and GPT-3.5.

The results of this review underscore that the main factor that affects performance, are the inherent capabilities of the LLM. Other factors, including various prompting methods, incorporation of questions that include media, and variability in question sets had secondary roles. This observation emphasizes the priority of advancing core AI model development to ensure better accuracy and utility in complex sectors like healthcare. As we consider the integration of LLMs into medical domains, it’s crucial to recognize that while optimizing external parameters can contribute to performance tweaks, the most profound improvements lie in the refinement of the model’s core capabilities.

The proficiency of these LLMs on a rigorous and foundational examination such as the USMLE provides a compelling indication of these models’ potential role in the medical domain. GPT-4’s ability to achieve high accuracy levels signifies the progression and maturation of AI’s capabilities in deciphering complex medical knowledge. Such advancements could be pivotal in assisting healthcare professionals, improving diagnostic accuracy, and facilitating medical education. However, while the results are promising, it is crucial to approach the integration of LLMs in medical practices with caution. The USMLE’s textual nature might not encompass the entire scope of clinical expertise, where skills like patient interaction,(19) hands-on procedures, and ethical considerations play an important role.

Prompting is considered to hold a significant role in shaping the performance of LLMs when answering queries.(20,21) This review demonstrates that the way questions are structured can subtly influence the responses generated by these models. Notably, open-ended prompting has a slight edge over the standard multiple-choice format. This suggests that LLMs might have a nuanced preference when processing information based on the context they’re provided. Moreover, the marginally better outcomes with 5-shot prompting compared to zero-shot hint at the LLMs’ capacity to adjust and produce informed answers when given a few guiding examples. Though these differences are subtle and may not dramatically change the overall performance, they provide valuable insights into the optimization of interactions with LLMs.

Two sets of formal USMLE questions were utilized in the studies reviewed. Both GPT-3.5 and GPT-4 exhibited superior performance on the Sample exam compared to the self-assessment. While the Sample exam is publicly accessible, the self-assessment can only be obtained through purchase. This raises the possibility that the higher accuracy is derived from previous encounters of the models with questions from the Sample exam. Nori et al.(17) developed an algorithm to detect potential signs of data leakage or memorization effects.

This algorithm is designed to ascertain if specific data was likely incorporated into a model’s training set. Notably, this method did not detect any evidence of training data memorization in the official USMLE datasets, which include both the self-assessment and the Sample exam. However, it is important to note that while the algorithm demonstrates high precision, its recall remains undetermined. Therefore, the extent to which these models might have been exposed to the questions during their training remains inconclusive.

In surveying the expansive literature on the application of LLMs in healthcare, it is noteworthy that only three studies are directly comparable in their use of formal question sets. This observation highlights the potential disparity in evaluation methods and emphasizes the need for standardized benchmarks in assessing LLMs’ medical proficiency. The USMLE stands as a primary metric for evaluating medical students and residents in the U.S., and its role as a benchmark for LLMs warrants careful consideration. While it offers a structured and recognized platform, it is crucial to contemplate whether such standardized tests can fully encapsulate the depth and breadth of LLMs’ capabilities in medical knowledge. Looking ahead there is a pressing need for research that delves into alternative testing mechanisms, ensuring a comprehensive and multidimensional evaluation of LLMs in the realm of healthcare.

This systematic review has several limitations. First, the studies reviewed primarily focused on multiple choice questions, which, although a prevalent format in the USMLE and an accepted method for assessing medical knowledge among students and clinicians, may not fully capture the complexity of real-world medical scenarios. Actual clinical cases often present with complexities and subtleties that might not strictly align with textbook descriptions. Hence, when contemplating the applicability of LLMs in a clinical setting, it is vital to recognize and account for this disparity. Secondly, our review intentionally excluded studies that examined other datasets used for USMLE preparation, such as MedQA. This decision was made to maintain consistency in the comparison of question sets. However, there is a wide array of research that evaluates various other LLMs using diverse question sets that mimic USMLE questions and measure medical proficiency. The exclusion of these studies potentially limits the comprehensiveness of our insights into the capabilities of LLMs in the medical domain.

To conclude, this systematic review presents a detailed analysis of the performance capabilities of LLMs on the USMLE. The primary influence of the core model’s capabilities over external factors, as highlighted in our findings, underscores the importance of continual advancements in AI model development targeted to the medical field. While the USMLE is a reputable metric for assessing medical knowledge, it might not wholly capture the diverse challenges of clinical practice. Future research should strive for a broader range of benchmarks, transcending traditional testing systems, to fully understand the potential and boundaries of LLMs in healthcare.

## Data Availability

All data produced in the present study are available upon reasonable request to the authors

